# Leptospirosis in Pregnancy: Prevalence, Risk Factors, Clinical Characteristics, and Outcomes in a North Indian Population

**DOI:** 10.1101/2022.11.02.22281830

**Authors:** Michal Shrestha, Saswati S Choudhury, Solomi V Carolin, Anjali Rani, Indrani Roy, Farzana Zahir, Swapna D Kakoty, Robin Medhi, Pranabika Mahanta, Rupanjali Deka, Manisha Nair

**Author notes:** Corresponding author (MN).

## Abstract

**Introduction:** Leptospirosis, a neglected zoonotic disease bears a significant global burden and is endemic in India. Leptospirosis in pregnancy can lead to severe adverse outcomes but is often misdiagnosed and underreported as clinical presentation is masked by the symptoms of common obstetric complications and other endemic infectious diseases. Our aim was to generate epidemiological information on the prevalence and distribution, risk factors, and outcomes (both maternal and fetal) of the disease among a population of pregnant and post-partum women in northern India.

**Methods:** We conducted a cross-sectional study on a random sample of 170 pregnant and post-partum women in 10 hospitals across three northern states of India in whom ELISA IgG and IgM antibody tests were done. Women were categorised as seropositive if they had high levels of either IgG or IgM. We identified study variables based on an available systematic review of leptospirosis in pregnancy. Fisher’s Exact and Wilcoxon rank-sum non-parametric tests were used to analyse the association between leptospirosis in pregnancy and a range of socio-demographic and clinical factors. An analysis of the geographical distribution of the seropositive cases was undertaken using Geographical Information System mapping.

**Results:** The prevalence of leptospirosis in pregnancy was 5.9% (95% CI: 2.85 – 10.55%) with 80% of the seropositive cases living near water sources and lowlands. Proportions of cardiac problems, raised liver enzymes, anaemia and adverse fetal outcomes were higher among women who were seropositive compared with women who were seronegative. Adverse maternal and fetal outcomes were more common among seropositive pregnant and post-partum women compared with seronegative women.

**Conclusions:** This is the first descriptive epidemiological study of leptospirosis in pregnancy in India that generates important hypotheses emphasizing the need for further research. Pregnant women in endemic regions should be screened for leptospirosis for early diagnosis and treatment to avoid adverse outcomes.

**Author Summary:** Leptospirosis bears a significant global burden and is endemic in India. In pregnancy, it can lead to serious complications, but leptospirosis in pregnancy is often misdiagnosed and underreported as clinical presentation of the disease is masked by the symptoms of common obstetric complications and other endemic infectious diseases. There are no detailed studies about leptospirosis in pregnancy in India with almost all published studies being case reports. In recognition of this limitation, this cross-sectional study was undertaken to understand the rates, risk factors and outcomes of leptospirosis in pregnancy in India. ELISA IgG and IgM antibody tests were conducted to determine the seroprevalence of leptospirosis in the population. The rate of leptospirosis in pregnancy in our study population was found to be 5.9%. A majority of the seropositive cases lived near water sources and lowlands. The adverse clinical presentation of cardiac problems, raised liver enzymes, anaemia and adverse fetal outcomes were observed to be more common among seropositive pregnant and post-partum women compared with seronegative women. The study highlights the need for more research and for screening of leptospirosis in pregnant women living in endemic regions for early diagnosis and treatment to avoid adverse outcomes.

## Introduction

Leptospirosis is a neglected, zoonotic, and waterborne disease, which infects more than a million people, and almost 60,000 deaths occur annually with 2.9 million Disability Adjusted Life Years (DALYs). It is endemic in tropical and subtropical regions of the world and is associated with poor income, low education, poor housing, absence of hygiene and sanitation [1–5]. South Asia’s tropical climate with high humidity provides an ideal environment for the survival of *Leptospira* pathogens making leptospirosis endemic in India [6]. Rodents and wide range of domestic and wild animals host and carry the pathogens which get transmitted to humans when they come in direct or indirect contact with urine from infected animals [7–9]. Leptospirosis is acquired through occupational exposure and are common among people living in rodent-infested, flood-prone or overcrowded urban areas.

Leptospirosis in pregnancy is underreported and is also often misdiagnosed as clinical presentation mimics symptoms of common obstetric complications such as Pre-Eclampsia, HELLP Syndrome (Haemolysis, Elevated Liver enzymes and Low Platelets) and Acute Fatty Liver of Pregnancy (AFLP) [10]. Presentation of leptospirosis is double masked in pregnancy, firstly due to the overlap in how the disease presents itself in the general population and in pregnant women and secondly because of the overlap between the signs, symptoms and physiological interferences associated with the disease and pregnancy complications mentioned above [5]. The prevalence of leptospirosis in pregnancy and its presentations and outcomes have not been extensively studied and there is paucity of epidemiological data. Systematic review conducted by Selvarajah et al. 2021 [5] to investigate the rates, risk factors and outcomes of leptospirosis in pregnancy estimated the incidence to be 1.3 per 10,000 women but also pointed out that this number could be higher in endemic areas. Leptospirosis in pregnancy, as in the general population, can present with fever, headache, muscle ache and jaundice which could escalate to adverse maternal and fetal outcomes of congenital infection, spontaneous miscarriage, abortion, maternal death, preterm birth, low birth weight in term birth, stillbirth, intrauterine fetal death (IUFD), neonatal death, preeclampsia, deranged liver functions, and respiratory problems [5]. The similarities of its signs and symptoms with pregnancy complications often leads to under- or mis-diagnosis of leptospirosis as it is not included in the immediate shortlist of differential diagnoses by health professionals. Further, the review suggested that adverse fetal outcomes were higher in pregnant women who were infected in early trimesters of pregnancy (first or second trimester) compared with women who were infected in their third trimester [5].

Even with subtle presentations, leptospirosis in pregnancy could be life-threatening and has adverse effect on both the mother and the fetus [10]. These adverse effect of leptospirosis to maternal and neonatal health is a serious concern. However, only 13% of published studies in leptospirosis from the WHO region of south-east Asia is on pregnancy according to Selvarajah et al [5]. There have not been any detailed epidemiological studies for leptospirosis in pregnancy in India with almost all published studies being case reports [5]. In recognition of this limitation, our aim was to conduct a cross-sectional study to understand the epidemiological context of leptospirosis in pregnancy in India with the following specific objectives:

1. To determine the seroprevalence of *Leptospira* antibody in pregnant women in the northern endemic region of India.
2. To geographically map the distribution of seropositive cases across the study region.
3. To compare the socio-economic and demographic characteristics of seropositive and seronegative pregnant/postpartum women.
4. To examine the association between maternal outcomes and *Leptospira* seropositivity.
5. To examine the association between fetal/infant outcomes and maternal *Leptospira* seropositivity.

## Methods

### Study-design and study settings

This was a cross-sectional study nested within an ongoing case-control study of Acute Heart Failure syndrome in pregnancy undertaken through the Maternal and perinatal Health Research collaboration, India (MaatHRI) [11]. The nested study was a random sample. It included the first 170 participants recruited consecutively in the main study in whom blood samples were tested for *Leptospira* antibody levels using enzyme-linked immunosorbent assay (ELISA) IgG (Immunoglobulin G) and IgM (Immunoglobulin M) antibody tests. Pregnant and postpartum (within 6 months of childbirth) women presenting with heart failure and control participants (women with no heart failure and giving birth immediately after the case) were recruited from ten hospitals across three northern States (Assam, Meghalaya, and Uttar Pradesh).

### Study variables

The selection of the study variables was informed by the systematic review of leptospirosis in pregnancy [5], and described in Table-1. Primary outcome was Leptospira seropositivity. Women having Leptospira IgM ≥ 1.1 Index and IgG ≥ 15 U/mL, measured using ELISA, were considered to have high seroprevalence of Leptospira antibodies, thus considered seropositive, based on the Indian population level cut-offs from the national reference laboratory. In the absence of standardised measures, the WHO recommends that population level values validated by local reference laboratories should be used to determine high seroprevalence [20]. Serum samples were analysed using ELISA to estimate levels of IgM (Kit – PANBIO) and IgG (Kit – DRG International, Inc). Serological diagnosis is considered to be the best method to detect *Leptospira* infections in humans. Specific antibodies appear in the blood around the fifth day after the onset of symptoms and detectable titres remain up to 10 days. Antibodies IgM usually appear early during the infection and before the IgG antibodies, therefore detection of IgM antibodies during the serological diagnosis is indicative of recent or beginning phase of the infection, whereas high titres of IgG antibodies indicate the recovery or convalescent phase of leptospirosis. IgG antibodies remain in the blood for years specifying history of past exposure to the disease. MAT (microscopic agglutination test) is considered as the gold standard diagnostic method for leptospirosis by the WHO (World Health Organization) but ELISA is commonly used as the test is more readily available in low resource settings, is user-friendly and has advantage of sensitivity and specificity over MAT [1,6,9,12,13].

**Table 1.** Description of the study variables.

| Variables of Interest |  | Definition |
| --- | --- | --- |
| Independent Variables | Dependent Variables |  |
| Seropositive cases for leptospirosis | | First the pregnant/postpartum women with antibodies IgM $\geq 1.1$ index and/ or IgG $\geq 15$ U/ml were identified. Separate variables for positive IgM and positive IgG were generated and then combined to generate 'seropositive cases', coded 1 for 'Yes' and 0 for 'No'. |
| Age |  | All women in the sample population were 18 years and above and up to 45 years of age. Age was used as a continuous variable. |
| Residence |  | Categorised as rural '1', urban '2' and suburb '3'. No women included in the study population lived in the suburbs. |
| Religion |  | Religion was categorised as '1' Hindu, '2' Muslim, '3' Christian, '4' Sikh and '5' Others. Due to small number of observations in each category, these were combined to generate a binary variable '1' Hindu and '2' Muslim, Christian and other minority religious communities. |
| Poverty |  | Below poverty line (BPL) was used to define poverty. The Planning Commission of India defines 'BPL' households as households (average 5 family members) with per capita consumption expenditure of INR 672.8 or less on a monthly basis in rural areas and INR 859.6 or less in urban areas at prices prevailing in 2009-10 (Planning Commission; Government of India, 2012).<br>Categorized as '1' BPL certificate/ self-certified, '2' Not BPL and '3' Not known. |
| Employment |  | Husbands who provided details of their occupation were categorized as '1' 'Employed' and without occupation as 0 'Unemployed'.<br>Women in paid employment 1 'Yes' and 2 'No'. |
| Education |  | Women's and husband's level of education was originally categorized into '1' Illiterate, '2' Literate upto 5th class, '3' 6th to 12th class, '4' Beyond 12th class, '99' Not known.<br>This was combined to generate a binary variable categorised as '1' Illiterate and primary and '2' Secondary and higher. |
| Smoking |  | Originally '1' Never, '2' gave up prior to pregnancy, '3' current and '4' gave up during pregnancy.<br>Combined to generate a binary variable '0' Never, '1' Current or in the past. |
| Tobacco consumption |  | Originally '1' Never, '2' gave up prior to pregnancy, '3' current and '4' gave up during pregnancy. |
|  |  | Combined to generate a binary variable '0' Never, '1' Current or in the past. |
| Alcohol consumption |  | Originally '1' Never, '2' gave up prior to pregnancy, '3' current and '4' gave up during pregnancy.<br>Combined to generate a binary variable '0' Never, '1' Current or in the past. |
| Chewing betel nut |  | Originally '1' Never, '2' gave up prior to pregnancy, '3' current and '4' gave up during pregnancy.<br>Combined to generate a binary variable '0' Never, '1' Current or in the past. |
| Consumes fermented fish |  | 1' Yes and 2 'No' |
|  | Cardio-vascular problems | Cardio-vascular problems is a composite variable comprising of women with cardiac problems and hypertensive disorders. '0' no cardio-vascular problem and '1' cardio-vascular problem/s present.<br>Cardiac problems included rheumatic heart disease, pulmonary oedema, dilated cardiomyopathy, myocarditis in shock, and peripartum cardiomyopathy.<br>Hypertensive disorders included eclampsia, pre-eclampsia, and pregnancy induced hypertension. |
|  | Deranged LFTs | Deranged LFTs (Liver Function Tests) was a composite variable for any deranged liver function parameters i.e. deranged AST/SGOT (Aspartate aminotransferase/ Serum glutamic-oxaloacetic transaminase), deranged ALT/ SGPT (Alanine transaminase/ Serum glutamic pyruvic transaminase), deranged GGTP (Gamma-glutamyl transpeptidase), deranged bilirubin, deranged ALP (Alkaline phosphatase) and deranged albumin. If the level of a parameter was higher than the recommended cut-off levels in pregnancy [14] it was considered as deranged coded as '1' and normal coded as '0':<br>• AST/SGOT >30 U/L was considered elevated and abnormal coded as '1' and <=30 coded '0',<br>• ALT/SGPT > 32 U/L was considered elevated and abnormal coded as '1' and <=32 coded '0',<br>• Bilirubin total > 14 mg/dL was considered elevated and abnormal coded as '1' and <=14 coded '0',<br>• GGTP > 41U/L was considered elevated and abnormal coded as '1' and <=41 coded '0',<br>• ALP > 418 U/L was considered elevated and abnormal coded as '1' and <=418 coded '0'.<br>• Albumin > 37 g/dL was considered elevated and abnormal coded as '1' and <=37 coded '0'.<br>Each parameter was also used as a continuous variable. |
|  | Respiratory problems | The variable 'respiratory problems' was coded '1' Yes and '0' No. Respiratory distress, tachypnea, shortness of breath, bronchial asthma, breathing difficulty were included. |
|  | Low antenatal haemoglobin / anaemia | Pregnant/ postpartum women who had haemoglobin level < 10g/dL during their first antenatal check-up were considered to have low haemoglobin and anaemic whereas those with >= 10g/dL were considered to have normal level of haemoglobin and non-anaemic [15]. |
|  | Maternal complications | Mortality, postpartum haemorrhage and sepsis were considered as maternal complications. |
|  |  | Mortality '1' woman died, '2' woman alive at six weeks postpartum follow-up, '99' not known<br>Postpartum haemorrhage – reported by the clinician<br>Sepsis – reported by the clinician |
| | Adverse infant or fetal outcomes | The composite variable adverse infant or fetal outcomes was constructed combining individual variables: stillbirth, admission to NICU, low birth weight (<2500gm) and premature gestational age at delivery.<br>Stillbirth was categorized as '0' Born alive and '1' Still born'.<br>Admission to NICU '1' for admitted to neonatal unit and '2' for not admitted.<br>Birth weight of babies born at term was considered low '1' if <2500 gm and '0' normal weight if $\geq 2500$gm.<br>Babies born at < 37 weeks' gestation were coded as '1' preterm birth and $\geq 37$ weeks' gestation as '0' term birth.<br>Infant mortality coded '1' for died and '2' for alive at 6 weeks postpartum follow-up. |

Determinants of leptospirosis in pregnancy [5], including demographic, economic and behavioural characteristics of pregnant women were considered as predictors or exposure variables for high seropositivity. Socio-demographic characteristics included age of the women, type of residence, religion, marital status. Socio-economic factors included poverty, education level of the women and their husbands, employment status of women and their husbands; and behavioural factors included smoking, consumption of tobacco and alcohol, chewing of betel nut and consuming fermented fish. These characteristics were compared between the seropositive and seronegative groups of pregnant women.

Clinical characteristics that have been identified as being associated with leptospirosis in pregnancy [5] such as cardio-vascular problems (cardiac complications and hypertensive disorders), deranged liver function, respiratory problems and low haemoglobin were compared between the two groups. Mortality and maternal complications such as postpartum haemorrhage and sepsis were also compared.

Adverse fetal and infant outcomes were analysed to compare their prevalence in the seropositive and seronegative groups of women. Information on pregnancy loss, stillbirth, low birth weight, premature birth, infant mortality and admission to NICU (Neonatal Intensive Care Unit) were used to generate a dummy variable for adverse fetal/infant outcomes. The analysis of fetal/infant outcomes was restricted to singleton pregnancies as the number of fetuses are likely to influence the selected adverse outcomes irrespective of leptospirosis infection.

### Methodology for Geographical Information System (GIS) mapping of the seropositive women

The geographical coordinates of residences of seropositive pregnant women were imported into the QGIS 3.16 software and mapped to analyse the environmental exposure of the seropositive pregnant/postpartum women [16]. The geographical coordinates were accessed with the help of Google maps using the participants’ residence addresses. The base layer and administrative boundary maps of India were obtained from *https://www.diva-gis.org/gdata* [17] and the CRS (coordinate reference system) used for mapping was EPSG:4326 WGS84 (World Geodetic System 1984). The digital elevation map (DEM) was a raster layer while other base layers were vector, and the data was imported to QGIS in a version of delimited text layer.

### Statistical Analysis

The *Leptospira* seropositivity rate with 95% CI (confidence interval) in pregnant/postpartum women in the study population was calculated as:

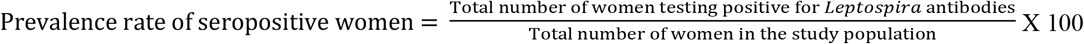

Categorical and binary independent variables were expressed as frequency and percentages and their proportions were compared between seropositive and seronegative groups. Fisher’s Exact test was used to examine whether the difference in proportions were statistically significant. Continuous independent variables were expressed as median with interquartile ranges and compared between the two groups. Wilcoxon rank-sum non-parametric test for difference in median was used to ascertain statistically significant difference in characteristics between the two groups.

All statistical tests were considered to be significant at a two-tailed p-value of 10%. The significance level was determined apriori to be 10% considering the small sample size. All analyses were performed using STATA version 17 [18].

### Patient and public engagement

The MaatHRI collaboration has a community engagement and involvement (CEI) group. The studies undertaken by the collaboration, including this study, receive regularly input and suggestions from the CEI group. The group is also involved in dissemination of the study outcomes.

## Results

As shown in the flow chart (Fig 1), the random study population of pregnant/postpartum women for whom *Leptospira* antibody tests was conducted was 170. Prevalence rate was calculated for the entire sample and GIS mapping was done for all seropositive participants.

**Fig 1.**
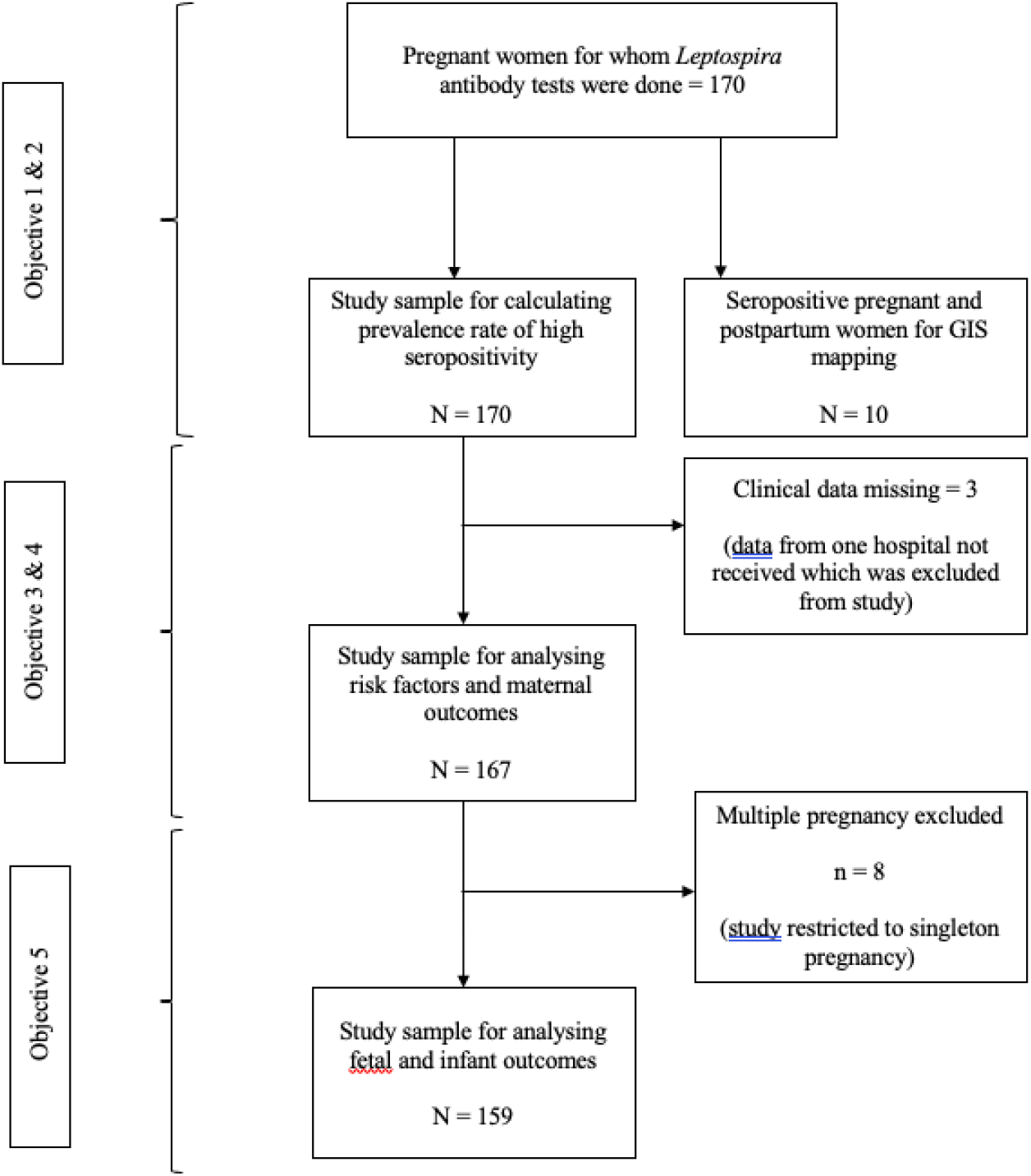
Flow chart of the study data.

Three participants did not have any clinical data and were therefore excluded from the other analyses. Analysis of fetal/infant complications was restricted to 159 singleton pregnancies (8 multiple pregnancies were excluded).

### Prevalence of leptospirosis and geographical mapping of seropositive cases in the study population in India

Among 170 women who underwent the IgM and IgG ELISA tests, 10 were determined as seropositive for *Leptospira* antibody. Thus, the overall calculated prevalence was 5.9%, 95% CI 2.85-10.55%, 4.11% for recent infection (n=7), 2.94% for previous infection (n=5) and two cases had high levels of both IgM and IgG antibodies.

Figure-2 presents the geographical mapping of the distribution of seropositive cases across northern India. From the plotted coordinates of the residences of seropositive women about 80% of the seropositive cases were observed to live near a river or natural water sources, specifically near the river-basins of the Brahmaputra river. The digital elevation map (DEM) in Figure-3 clearly demonstrates the seropositive cases inhabiting the river-basins and the lowlands.

**Fig 2.**
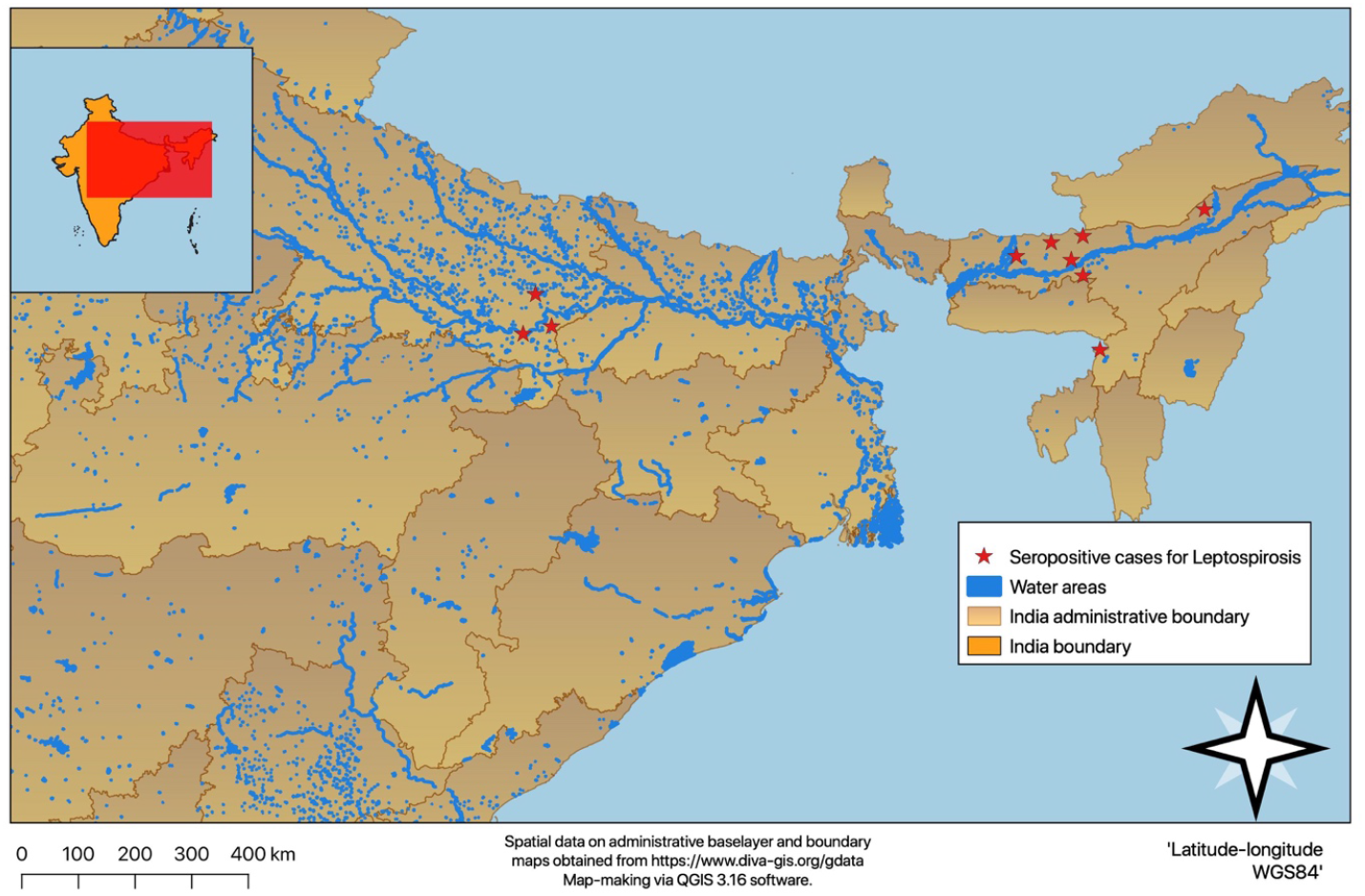
Geographical distribution of seropositive cases for leptospirosis infection.

**Fig 3.**
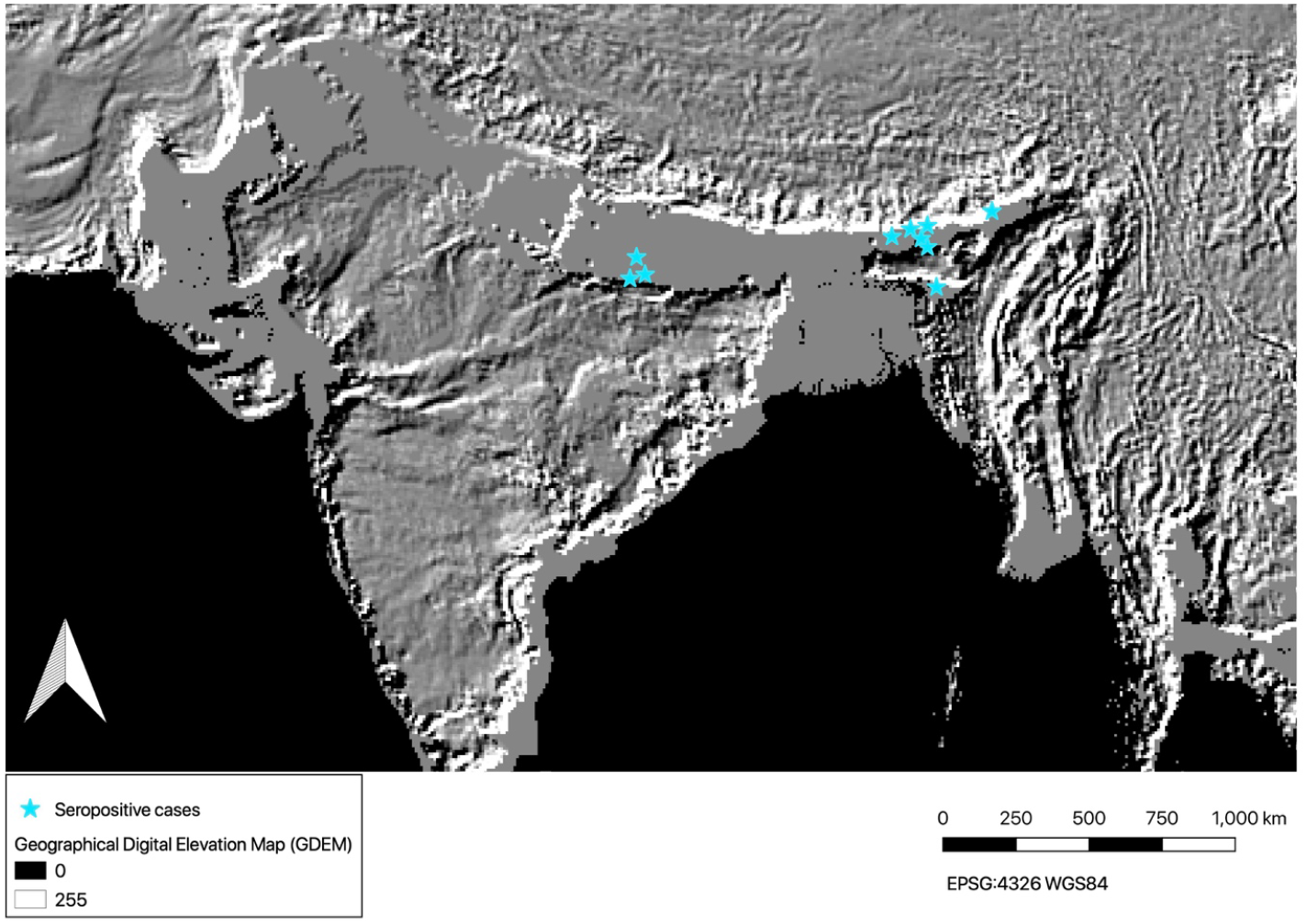
Distribution of seropositive cases for leptospirosis on a digital elevation map.

### Socioeconomic, demographic, and behavioural characteristics

None of the socioeconomic, demographic or behavioural characteristics were found to be statistically significantly different between the seropositive and seronegative groups (Table-2) but there were marked differences in proportion of some factors. Majority of seropositive women i.e., 90% lived in rural areas whereas this was 75.8% in the seronegative group, and 80% of the seropositive group followed Hinduism. Median age of women was comparable for both groups. None of the women in the study population were reported to smoke or consume alcohol or use tobacco. The proportion of women who currently chewed or had a history of chewing betel nut was 50% for the seropositive group which was higher than 33.1% in the seronegative group, although not statistically significant.

**Table 2.** Socioeconomic, demographic and behavioral characteristics of the study population.

| Total sample of pregnant women (N) = 167 |  |  |  |  |
| --- | --- | --- | --- | --- |
| Socioeconomic, demographic & behavioural characteristics: |  | Seropositive women | Seronegative women | P-value* |
|  |  | n = 10 | n = 157 |  |
|  |  | n (%) |  |  |
| Residence |  |  |  | 0.46 |
|  | Rural | 9 (90.0 %) | 119 (75.8 %) |  |
|  | Urban | 1 (10.0%) | 38 (24.2 %) |  |
| Religion |  |  |  | 0.50 |
|  | Hindu | 8 (80.0%) | 102 (65.0%) |  |
|  | Muslim, Christian & others | 2 (20.0%) | 55 (35.0%) |  |
| Poor |  |  |  | 0.44 |
|  | BPL | 4 (40.0%) | 90 (57.3%) |  |
|  | not BPL | 5 (50.0%) | 55 (35.0%) |  |
|  | not known | 1 (10.0%) | 12 (7.6) |  |
| Women's education level |  |  |  | 0.69 |
|  | Illiterate & primary | 2 (20.0%) | 27 (17.2%) |  |
|  | Secondary & higher | 8 (80.0%) | 130 (82.8%) |  |
| Husband's education level |  |  |  | 0.21 |
|  | Illiterate & primary | 0 (0.0%) | 30 (19.1%) |  |
|  | Secondary & higher | 10 (100.0%) | 127 (80.9%) |  |
| Chewing betel nut |  |  |  | 0.31 |
|  | Current or has history | 5 (50.0%) | 52 (33.1%) |  |
|  | Never | 5 (50.0%) | 105 (66.9%) |  |
| Consumes fermented fish |  |  |  | 0.29 |
|  | Yes | 5 (50.0%) | 109 (69.4%) |  |
|  | No | 5 (50.0%) | 48 (30.6%) |  |
|  |  | **Median (IQR)** | **Median (IQR)** | **P-value\*\*** |
| Age (in years) |  | 25 (5) | 24 (6) | 0.71 |

\* P-value Fishers Exact test for categorical variables
\*\* P-value Rank-sum test for continuous variable
P-value < 0.10 (10% significance level)
IQR – Interquartile range; BPL – below poverty line

### Clinical Characteristics

Clinical characteristics of the seropositive and seronegative women are presented in Table-3. A significantly higher proportion (70%) of the seropositive women were reported to have cardio-vascular problems compared with 32.5% in the seronegative group (p-value of 0.03). On further examining the individual components of the composite variable, a higher proportion of seropositive women had co-existing cardiac problems (60%) compared with 47% in the seronegative group (p-value 0.08). Similarly, proportion of co-existing hypertensive disorders of pregnancy was higher in the seropositive group (40%) compared with the seronegative group (16%) at a p-value of 0.07.

**Table 3.** Clinical characteristics of the study population.

| Total sample of pregnant women (N) = 167 |  |  |  |  |
| --- | --- | --- | --- | --- |
| Clinical characteristics: |  | Seropositive women | Seronegative women | P-value* |
|  |  | n = 10 | n = 157 |  |
|  |  | n (%) | n (%) |  |
| **Cardio-vascular problems ‡** |  |  |  | 0.03 |
|  | Yes | 7 (70.0%) | 51 (32.5%) |  |
|  | No | 3 (30.0%) | 106 (67.5%) |  |
| **Cardiac problems** |  |  |  | 0.08 |
|  | Yes | 6 (60.0%) | 47 (29.9%) |  |
|  | No | 4 (40.0%) | 110 (70.1%) |  |
| **Hypertensive disorders in pregnancy** |  |  |  | 0.07 |
|  | Yes | 4 (40.0%) | 25 (15.9%) |  |
|  | No | 6 (60.0%) | 132 (84.1%) |  |
| **Deranged LFTs ‡** |  |  |  | 0.5 |
|  | Yes | 8 (80.0%) | 100 (63.7%) |  |
|  | No | 2 (20.0%) | 57 (36.3%) |  |
| **Deranged AST** |  |  |  | 0.2 |
|  | Yes | 8 (80.0%) | 90 (57.3%) |  |
|  | No | 2 (20.0%) | 67 (42.7%) |  |
| **Deranged ALT** |  |  |  | 0.28 |
|  | Yes | 4 (40.0%) | 39 (24.8%) |  |
|  | No | 6 (60.0%) | 118 (75.2%) |  |
| **Deranged GGTP** |  |  |  | 0.32 |
|  | Yes | 2 (20.0%) | 17 (10.8%) |  |
|  | No | 8 (80.0%) | 140 (89.2%) |  |
| **Anaemia** |  |  |  | 0.09 |
| (Hb < 10g / dL) | Yes | 7 (70.0%) | 58 (37.9%) |  |
|  | No | 3 (30.0%) | 95 (62.1%) |  |
|  | missing |  | 4 |  |
| **Respiratory Problems** |  |  |  | 0.59 |
|  | Yes | 1 (10.0%) | 13 (8.3%) |  |
|  | No | 9 (90.0%) | 144 (91.7%) |  |
|  |  | **Median (IQR)** | **Median (IQR)** | **P-value\*\*** |
| **Liver Function Parameters:** |  |  |  |  |
|  | ASTSGOT (U/L) | 48.5 (26) | 37 (44) | 0.29 |
|  | ALTSGPT (U/L) | 21.5 (36) | 18 (21) | 0.49 |
|  | GGTP (U/L) | 21 (25) | 17 (20) | 0.53 |
|  | ALP (U/L) | 222.5 (112) | 209 (121) | 0.93 |
|  | Bilirubin (mg/dL) | 0.195 (0.1) | 0.2 (0.11) | 0.94 |
|  | Albumin (g/dL) | 2.68 (0.68) | 2.7 (0.82) | 0.97 |
| **Haemoglobin in g/dl** |  |  |  |  |
|  | Antenatal Haemoglobin level | 9.2 (2) | 10.2 (2) | 0.04 |
|  | missing |  | 4 |  |

\* P-value Fishers Exact test for categorical variables
\*\* P-value Rank-sum test for continuous variable
P-value < 0.10 (10% significance level)
‡ Composite variable
Missing data were not included in the calculation of percentages and median
IQR – interquartile range

Although not significantly different at p<0.1, the proportion of women with deranged LFT (Liver Function Test) was higher in the seropositive group (80%) compared with 64% in the seronegative group. The median levels of all individual parameters included in the composite deranged LFT variable were found to be elevated in the seropositive group compared with the seronegative group, but these were not statistically significantly different. The median antenatal Hb (Haemoglobin) level was 9.2 g/dl in seropositive group of pregnant and postpartum women compared with 10.2 g/dl in women who were seronegative (p=0.04), thus a higher proportion of the seropositive women had antenatal anaemia compared with the seronegative group. Respiratory problems were reported in 10% of the seropositive women and 8.3% among seronegative women (p = 0.59).

None of the women in the seropositive group died but 3 maternal deaths were reported in the seronegative group and 1 from seronegative group reported to have sepsis. No woman was reported to have a postpartum haemorrhage.

### Fetal and infant outcomes

The proportion of adverse infant/fetal outcomes was significantly higher in the seropositive group compared with the seronegative group at p-value of 0.08 (Table-4). There was one loss of pregnancy in the seropositive group, the reason for which was not known, and no infant died among women with singleton pregnancies.

**Table 4.** Infant and fetal outcomes of the study population restricting to singleton pregnancies.

| Total sample of pregnant women (N) = 159 |  |  |  |  |
| --- | --- | --- | --- | --- |
| Infant and fetal outcomes |  | Seropositive women | Seronegative women | P-value* |
|  |  | n = 9 | n = 150 |  |
|  |  | n (%) |  |  |
| **Adverse infant/fetal outcomes ‡** |  |  |  | 0.08 |
|  | Yes | 7 (87.5%) | 78 (54.2%) |  |
|  | No | 1 (12.5%) | 66 (45.8%) |  |
|  | missing | 1 | 6 |  |
| **Stillbirth** |  |  |  |  |
|  | Yes | 1 (12.5%) | 5 (3.5%) | 0.28 |
|  | No | 7 (87.5%) | 139 (96.5%) |  |
|  | missing | 1 | 6 |  |
| **Admission to NICU** |  |  |  | 0.03 |
|  | Yes | 4 (57.1%) | 24 (17.3%) |  |
|  | No | 3 (42.9%) | 115 (82.7%) |  |
|  | missing | 2 | 11 |  |
| Low birth weight (< 2500 gm) |  |  |  | 1.00 |
|  | Yes | 4 (50.0%) | 64 (44.4%) |  |
|  | No | 4 (50.0%) | 80 (55.6%) |  |
|  | missing | 1 | 6 |  |
| Preterm birth (<37 weeks' gestation) |  |  |  | 0.22 |
|  | Yes | 4 (50.0%) | 39 (27.1%) |  |
|  | No | 4 (50.0%) | 105 (72.9%) |  |
|  | missing | 1 | 6 |  |
|  |  | **Median (IQR)** | **Median (IQR)** | **P-value\*\*** |
| Median birth weight |  | 2455.5 (882.5) | 2600 (649) | 0.28 |
|  | missing | 1 | 6 |  |
| Median gestational age at delivery |  | 36.4 (3.4) | 39 (3.9) | 0.12 |
|  | missing | 1 | 6 |  |

\* P-value Fishers exact for categorical
\*\* P-value Rank-sum for continuous variable
P-value < 0.10 (10% significance level)
‡ Composite variable
Missing data were not included in the calculation of percentages and median

A summary of the clinical characteristics, co-morbidities, maternal and fetal outcomes of the seropositive women are presented in Table-5. IgM was higher in seven women suggesting acute infection and three women had high IgG and can be considered to be in convalescent phase. A majority of the women had high AST (8 out of 10) and low haemoglobin (7 of 10). Comorbidities were reported as cardiac problems in six women and hypertensive disorders in pregnancy in four. There was one case of stillbirth and one pregnancy loss due to miscarriage. Caesarean section was performed on three women and only one had a twin pregnancy. Low birth weight, premature birth and admission of the baby to NICU was common among women who were categorised as seropositive in the study population. Seven out of ten seropositive women had at least one adverse fetal/infant outcome.

**Table 5.** Summary of clinical presentation, comorbidities, maternal and fetal outcomes.

| Seropositive cases* | Blood Parameters |  |  |  |  |  | Co-morbidities |  | Pregnancy Outcomes |  |  |  |  |  |  |  | Infant Outcomes |  |  |
| --- | --- | --- | --- | --- | --- | --- | --- | --- | --- | --- | --- | --- | --- | --- | --- | --- | --- | --- | --- |
|  | IgG value | IgM value | High AST | High ALT | High GGTP | Hb at first antenatal visit (<10g/dl) | Diagnosed hypertensive disorders in pregnancy | Reported cardiac problems | Multiple pregnancy | Caesarean section | Miscarriage | Stillbirth | Mother died | Post-partum haemorrhage | Multi-organ failure | Respiratory problems | Birth weight of infant (<2500g) | Premature birth (<37wks) | Admission to NICU |
| SP-1 | 15 | 0.53 |  |  |  |  |  | X |  |  |  |  |  |  |  |  | X | X | X |
| SP-2 | >100 | 0.8 | X |  |  | X | X | X |  | X |  |  |  |  |  | X |  | X |  |
| SP-3 | 25 | 1.1 |  |  |  | X | X | X |  |  | X |  |  |  |  |  |  |  |  |
| SP-4 | 6 | 1.34 | X | X |  | X |  | X |  | X |  |  |  |  |  | X |  |  | X |
| SP-5 | 15 | 1.2 | X |  |  | X |  |  |  |  |  |  |  |  |  |  | X |  |  |
| SP-6 | 2.6 | 1.14 | X |  |  | X | X |  | X |  |  |  |  |  |  |  | X | X | X |
| SP-7 | 3 | 1.12 | X |  |  |  | X | X |  | X |  |  |  |  |  |  | X | X | X |
| SP-8 | 2.4 | 1.23 | X | X | X | X |  | X |  |  |  |  |  |  |  |  |  |  |  |
| SP-9 | 4 | 1.32 | X | X |  | X |  |  |  |  |  |  |  |  |  |  | X | X | X |
| SP-10 | 16 | 0.33 | X | X | X |  |  |  |  |  |  | X |  |  |  |  |  |  |  |

\* If either IgG ≥ 15 or IgM ≥ 1.1
SP = Women who had seropositive test for leptospirosis
X = Occurrence of event

## Discussion

### Summary of the findings

The study estimated a prevalence rate of 5.9% for leptospirosis in pregnancy among a north Indian population. GIS mapping showed that the distribution of majority of seropositive cases were along the major river Brahmaputra and in the river-basin. Socio-economic factors were not found to be significantly associated with seropositivity in the study population, but this could be due to the small sample size. Adverse infant outcomes were common among women who were seropositive. There was one reported miscarriage and one stillbirth in the seropositive group, but this number could be higher in a larger population. Admission of infant to NICU was more frequent for the seropositive mothers compared with the seronegative mothers. LFTs were raised for the seropositive women, and they were also diagnosed with antenatal anaemia and cardiovascular problems.

### Prevalence of leptospirosis in pregnancy in northern India

Among the 170 randomly selected pregnant women for whom the IgM and IgG ELISA tests for *Leptospira* antibodies was performed, 10 tested positive giving a prevalence rate of 5.9% which is high if we consider the pooled rate of 1.3% from 17 countries reported by Selvarajah et al [5] in a systematic review of leptospirosis in pregnancy. The review however indicated that the number could be higher in endemic regions considering that the incidence rate in a study from Mexico was reported to be 13.6% [5,19].

### Diagnosis of leptospirosis in pregnancy

India is an endemic area for leptospirosis infection and as per WHO recommendation, the local laboratory cut-offs for titres of antibodies IgM and IgG were used to decide on high seroprevalence [20]. There are several diagnostic methods for testing seroprevalence of antibodies such as MAT (microscopic agglutination test); culture of blood, urine, tissues, and cerebrospinal fluid; polymerase chain reaction (PCR) and ELISA. Leptospirosis is confirmed by an increase in IgM antibody titres in the acute or initial phase of the disease indicating a current or recent infection [1,9,12,21]. However, the IgM titres produced in the first week of infection can remain for months to years raising question on whether its detection confirms acute or recent infection. WHO recommends MAT as a confirmatory test after detection of antibodies for leptospirosis through ELISA, which was not done in our study population. Nevertheless, ELISA is considered the best pragmatic method as it is a rapid test, is easily available, has better sensitivity and specificity and does not require expert knowledge in handling samples and performing tests. In pregnancy, leptospirosis infection could have serious consequences for both the mother and the fetus if not diagnosed early and left untreated [5]. Therefore, early detection through ELISA could be an affordable and pragmatic screening test for women suspected to have an infection in the endemic areas. However, the challenge currently seems to be non-inclusion of leptospirosis as a differential diagnosis for women who present with features of AFLP and HELLP even in endemic areas [5].

### Geographical distribution of the leptospirosis cases in the study region

The majority of the women who tested positive were found to live in the river basin of a large river called ‘Brahmaputra’ in the Northeastern part of India where flood hazard is a major problem. This region is also famous for having the heaviest rainfall in the world [22,23]. Rain washes off the topsoil during floods creating neutral and alkaline soil which facilitates the growth and survival of *Leptospires* in the environment. The *Leptospires* thrive in fresh water and in moist alkaline soil for weeks to years increasing the rate of transmission among both humans and animals. The groundwater used for drinking and other daily activities in the region also come from the same water table exposing the residents to the bacteria. Leptospirosis is well-known to be a water-borne disease but the bioburden of pathogens in the soil is often underestimated, which transmits to humans via skin [1]. The geospatial distribution in our study population suggests that environmental exposure could be one of the vital risk factors for leptospirosis among the study population.

### Risk factors for leptospirosis in pregnancy

Several studies conducted in low- and middle-income countries (LMICs) have found socioeconomic and demographic risk factors to be associated with leptospirosis, but in our study, these characteristics were not different between the seropositive and seronegative pregnant and postpartum women. This could be due to small sample size of the study. Pregnant and postpartum women residing in the rural areas and from Hindu religious background appeared to be more prone to the infection, but this could be because a majority of the women in the study area lived in rural regions and followed Hinduism. Smoking, consuming alcohol and tobacco, chewing betel nut were also not found to be higher among seropositive women. There was also no association between hygiene and seropositivity which is in contrast to Cook et al’s [21] study that found leptospirosis to be related to personal hygiene factors due to a greater risk of transmission.

### Clinical characteristics of leptospirosis in the study population of pregnant and postpartum women

Frequency of cardiac involvement in leptospirosis is underreported and often leptospirosis is not suspected to be a cause for cardiovascular complications as various other factors could induce this. Studies show mortality resulting from acute renal failure, acute respiratory distress syndrome and pulmonary haemorrhage to be associated with leptospirosis, but heart failure is also observed in severe cases of leptospirosis and there is evidence to support this in post-mortem studies of patients who died due to leptospirosis [24]. A study on leptospirosis outbreak in 2008 in Sri Lanka reported patients with heart failure and myocarditis [25]. In our study, prevalence of cardiovascular problems including cardiac problems and hypertensive disorders in pregnancy were observed to be higher in seropositive women compared with seronegative women. Further, the median level of AST from the liver function tests was higher in the seropositive cases and a majority of the women who had elevated levels of AST also had elevated levels of IgM antibodies. This might be an indicator of an acute infection phase of leptospirosis. Haemoglobin level less than 10g/dL was observed in 70% of the women who tested positive for leptospirosis which is consistent with the findings from a study by Edmond Puca et al. who found haemolytic anaemia to be associated with leptospirosis infection in humans [26]. Although iron deficiency and haemoglobinopathies were the main cause of anaemia in our study population, we do not know whether any of the participants had haemolytic anaemia. Studies have reported pulmonary haemorrhage and respiratory distress syndrome as an outcome of leptospirosis, but this was not observed in our study population. As reported in the systematic review of leptospirosis in pregnancy [5] and other studies [27,28], a significantly higher proportion of seropositive women in our study population had adverse fetal outcomes compared with the seronegative women. Leptospirosis in pregnancy resulted in lower median birth weight and preterm delivery. NICU admissions of infants was more frequent for seropositive mothers compared with seronegative mothers. There was one reported miscarriage and one stillbirth, but maternal death or complications such as postpartum haemorrhage was not observed to be higher among seropositive women.

### Strengths and limitations

This is the first study in India describing the epidemiology of leptospirosis among pregnant and postpartum women. Geographical mapping and analyses of the data generates important hypotheses about the distribution of the cases, transmission risk, clinical characteristics, and outcomes of leptospirosis in pregnancy suggesting the urgent need for further research. A major limitation of this study is the small sample size which limited the study power and therefore the ability to undertake further statistical analyses. However, it is important to acknowledge that leptospirosis in pregnancy is not common and a majority of the studies published till date are case reports or case series. Furthermore, although ELISA tests are antibody specific, the risk of contamination or high titres due to other infections cannot be ignored. Infections such as malaria, dengue or other unknown infections could have influenced the ELISA detected antibodies [29], although the chances of this is low as there was no reported infections in the clinical data collected from the participants’ hospital records.

## Conclusions

Leptospirosis has been recognised as a Neglected Zoonotic Disease (NZD) by the WHO as this affects the livelihoods of those infected along with their health, perpetuating poverty [30]. Despite being an NZD and given the complex epidemiological pattern of the disease, there still remains a lack of focus on policies and strategies for prevention and control of leptospirosis and the global burden is high [31]. This hypothesis generating study showed that the prevalence rate of leptospirosis in pregnancy could be higher than that estimated in previous studies and therefore there is a requirement for more research and epidemiological surveillance of leptospirosis in pregnancy and in the general population. ELISA tests for IgM and IgG antibodies could be used to screen pregnant women during their routine antenatal check-ups in endemic regions, particularly those who present with non-differential febrile illness. Pregnant women presenting with high blood pressure are diagnosed as having hypertensive disorders of pregnancy with leptospirosis not being suspected as a differential diagnosis. This study suggests that in endemic areas it is important to investigate leptospirosis infection as a differential diagnosis to initiate appropriate treatment to avoid adverse pregnancy and fetal outcomes. Even though our study did not find socio-economic characteristics to be significantly different between the seropositive and seronegative groups, likely due to a small sample size, clinicians and public health professionals should not ignore the role played by these factors in increasing the risk of leptospirosis. Leptospirosis in pregnancy can be prevented and clinically managed through proper awareness and education of the general population and health professionals. However, a key first step is to include leptospirosis in the list of differential diagnosis of febrile illness and hypertensive disorders during pregnancy.

## Data Availability

All data available for this study are included in the paper and can also be obtained for free. Information on how to access data is available on Oxford Population Health Department’s website (https://www.ndph.ox.ac.uk/data-access) and requests can also be emailed directly to.

## Acknowledgements

We thank the study participants for their willingness to participate in the study. We thank the MaatHRI field team for collecting the data. We also thank Dr Sinaida Cherubin, who provided help to the first author in understanding the data.

